# Protocol for disease-oriented Russian disc degeneration study (RuDDS) biobank facilitating functional omics studies of lumbar disc degeneration

**DOI:** 10.1101/2021.03.07.21253070

**Authors:** Olga N. Leonova, Elizaveta E. Elgaeva, Tatiana S. Golubeva, Alexey V. Peleganchuk, Aleksandr V. Krutko, Yurii S. Aulchenko, Yakov A. Tsepilov

## Abstract

**Introduction:** Lumbar intervertebral disc degeneration (DD) disease is one of the main risk factors for low back pain. The social and economic importance of low back pain is very high: back pain is among the leading causes of absenteeism and the cost of treating back pain exceeds the cost of treatment of many other serious diseases (cancer, in particular); however, therapy does not always provide the desired result. Despite the variability of biological studies of lumbar DD, it is still not fully understood, partially due to the fact that there are only few studies using systematic and integrative approaches. Hence, more integrative omics studies are needed to link all pieces of knowledge together, build a complete picture of biology of lumbar DD and obtain a deeper understanding of the processes underlying this pathology.

**Methods and analysis:** This disease-oriented biobank to study lumbar disc degeneration will be recruited from two clinical centers. A total of 1100 participants with available lumbar MRI will be enrolled during the three-year period. General information about a patient, medical history, lumbar MRI parameters and biological material (whole blood and plasma) will be collected in the centers at baseline. Then, from those patients, who will undergo a spine surgery during the treatment, disc tissue samples will be gained. Eventually, postoperative clinical data will be collected from operated patients during the follow-up.

**Ethics and dissemination:** The study will be performed according to the Helsinki Declaration. The study protocol was approved by the Local Ethical Committee of NRITO (№034/20 dated 02 Oct 2020) and by the Local Ethical Committee of Priorov CITO (№1/21 dated 25 Feb 2021). Preliminary and final results will be presented in peer-reviewed journals, at national and international congresses.

**Trial registration number:** NCT04600544

**ARTICLE SUMMARY:** *STRENGTHS AND LIMITATIONS OF THIS STUDY:*

- Two-center and multiple-discipline study: the study includes two centers (Moscow in the European and Novosibirsk in the Asian part of Russia), with research teams specializing in following fields: (1) clinical aspects of lumbar disc degeneration disease, (2) biology and genetics of pain, (3) generation of omics data and (4) multi-omics data analysis
- Collection of variable biological material: whole blood, plasma and, for part of the participants, intraoperative material of the lumbar disc (with different DD status) will be available for omics studies
- Objective diagnosis: lumbar DD status is confirmed by MRI, not self-reported diagnosis
- Sample size is limited compared to nationwide biobanks
- Patients with severe lumbar DD status will be more prevalent in the biobank than these with mild or no lumbar DD compared to the general population

## INTRODUCTION

Intervertebral disc degeneration is a normal aging process, but in some cases it causes lumbar disc degeneration disease (LDDD) [1,2]. Intervertebral disc degeneration (DD) often begins earlier than degenerative changes in the ligaments, cartilages and other tissues of spinal segment [3]. DD is a major contributor to low back pain [4], but also DD can proceed without back pain [5–8]. However, LDDD is associated with lower health-related quality of life [9] and is the leading cause of population absenteeism and disability [10,11].

The prevalence of lumbar DD in the general population is extremely high: up to 50% in people of 30-39 years old [2,12]. There is not a single adult without even a small degree of degeneration in the intervertebral discs [13].

List of well-known risk factors of LDDD includes female gender, advanced age [14], obesity, smoking [15], absence of or extreme physical activity [16] and genetic risk factors [17]. However, even in the absence of obvious risk factors, there are cases of LDDD among young people [1,14], as well as progressive severe LDDD requiring a number of surgical interventions.

Biology of intervertebral DD is actively studied from different points of view. Besides various studies of degenerative disc morphology [18–22] and molecular biology [16,23–25] there is an increasing number of genetic [26,27], transcriptomic [28–30] and proteomic [31–33] studies of intervertebral DD. Thus, nowadays based on candidate-gene [34] and genome-wide associated studies (GWAS) [35,36] over 160 genes were considered as potentially involved in intervertebral disc degeneration, although less than ten of them provide strong evidence for the association [34]. More than 500 genes were highlighted as differentially expressed in degenerative disc tissue in comparison with healthy intervertebral discs [37,38]. Moreover, proteomic [31–33] and metabolomics [39] changes were also detected in degenerative intervertebral discs. However, despite a large number of studies of LDDD, it is still not fully understood, partially due to the fact that there are only few studies using systematic and integrative approaches [40]. Hence, more integrative omics studies are needed to link all pieces of knowledge together, build a complete picture of biology of lumbar DD and obtain a deeper understanding of the processes underlying this pathology.

### Research aim and objectives

The main aim of this study is to establish disease-oriented biobank to facilitate research in biology of the lumbar disc degeneration. Diverse biological samples (whole blood, plasma, disc tissue) along with MRI imaging, clinical, socio-demographic and various omics data (e.g. genomic and transcriptomic) will be available for researchers and clinicians for a variety of further multi-omics studies. It will lay the groundwork for the development of early diagnostics of DDD and its personalized treatment.

## METHODS AND ANALYSIS

### Patient and Public Involvement

Patients and/or the public were not involved in the design, or conduct, or reporting, or dissemination plans of this research.

### Study design and settings

This disease-oriented biobank to study lumbar disc degeneration will be recruited from two centers: Priorov National Medical Research Center of Traumatology and Orthopedics (Priorov CITO), Moscow, Russia and Novosibirsk Research Institute of Traumatology and Orthopedics (NRITO), Novosibirsk, Russia. It will include only patients with available MRI of the lumbar spine, who signed the informed consent. Clinical data and specimen collection will be performed in three visits as described in Table 1. At the first step general information about a patient, his medical history and MRI scans of lumbar spine will be obtained. Patient’s biological material (whole blood and plasma) will be also sampled in the centers at baseline. Then, from those patients, who will undergo a spine surgery during the treatment, disc tissue samples will be gained. Eventually, postoperative clinical data will be collected from operated patients during the follow-up. During three years, we expect to collect information and biological material from a total of 1100. The full scheme of the study is presented at Figure 1.

**Table 1.** Scheduled procedures for clinical data and samples collection.

|  | Visit 1 - Baseline | Visit 2 - Day of surgery<sup>1</sup> | Visit 3 - 3 months after surgery<sup>2</sup> |
| --- | --- | --- | --- |
| Visit Window (±Days) |  |  | Day 90 ±14 days |
| Informed Consent | X |  |  |
| Eligibility Criteria | X |  |  |
| Medical History | X |  |  |
| Anamnesis Vitae | X |  |  |
| Demographics | X |  |  |
| Blood samples | X |  |  |
| MRI | X |  |  |
| ODI<sup>3</sup> | X |  | X |
| VAS<sup>4</sup> | X |  | X |
| Indications for surgery |  | X |  |
| Surgical Procedure |  | X |  |
| Intervertebral disc samples |  | X |  |
| Adverse events / Serious adverse events | X | X | X |

<sup>1</sup>only for patients subjected for surgery, <sup>2</sup>optional, <sup>3</sup>Oswestry Disability Index (ODI) scale, <sup>4</sup>Visual analogue scale (VAS) for back and leg pain.

**Figure 1.**
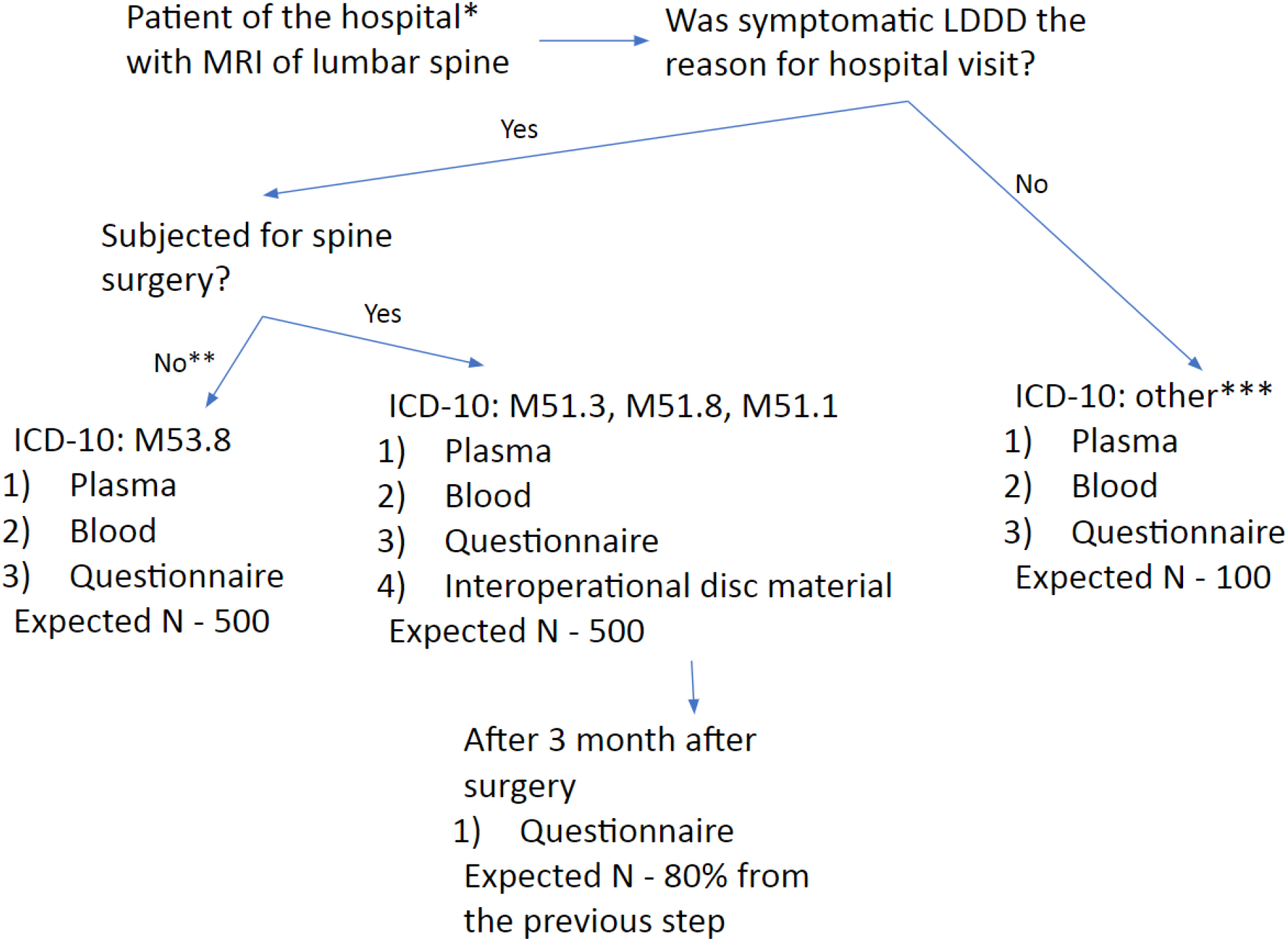
Patient selection. Sample size is depicted for 3 years timeline of project duration. Expected total sample size is 1100. *Novosibirsk Research Institute of Traumatology and Orthopedics (NRITO) / Priorov National Medical Research Center of Traumatology and Orthopedics (Priorov CITO). **Conservative treatment. ***ICD-10: I71 Aortic aneurysm and dissection, K81 Cholecystitis, K85 Pancreatitis, K25, K26 Penetrating ulcer, M06.4 Inflammatory polyarthropathy, N00-N99 Diseases of the genitourinary system

All biological samples from NRITO will be then transported to the Institute of Cytology and Genetics (ICG), Novosibirsk, Russia for long-term storage, further processing and omics profiling, including genotyping of blood samples and RNA-Sequencing of disc tissue. Priorov CITO will store the specimens on its own.

### Patient selection

Participants aged over 18 with available MRI scans of lumbar spine, who will also sign an informed consent and meet all the eligibility criteria (Table 2), will be included in the study. All participants will be grouped by three groups based on their diagnosis and type of treatment (conservative or surgical). All details and ICD-10 codes required for inclusion are presented in Figure 1. Restrictions on participation in the study for patients with bloodborne pathogens is determined by requirements of laboratory biosafety.

**Table 2.**
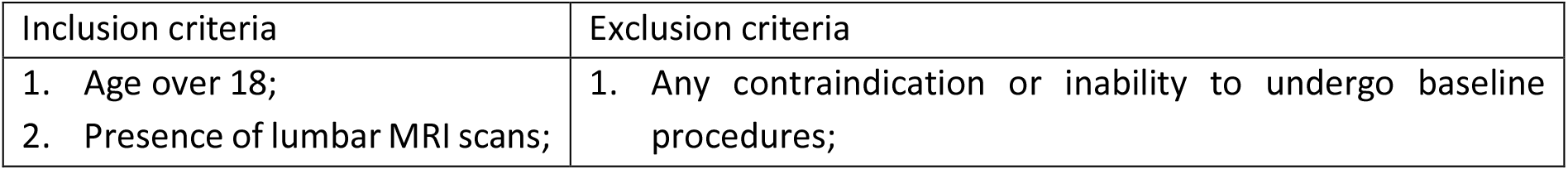
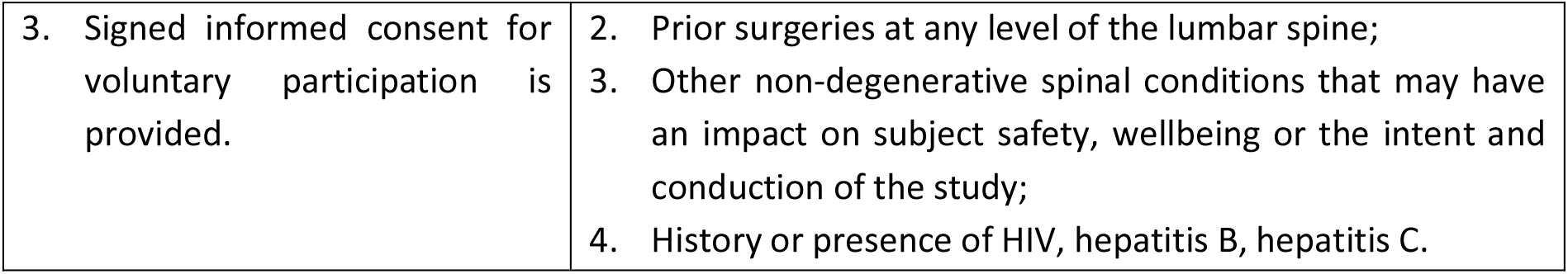
Eligibility criteria.

| Inclusion criteria | Exclusion criteria |
| --- | --- |
| 1. Age over 18;<br>2. Presence of lumbar MRI scans; | 1. Any contraindication or inability to undergo baseline procedures; |
| 3. Signed informed consent for voluntary participation is provided. | 2. Prior surgeries at any level of the lumbar spine;<br>3. Other non-degenerative spinal conditions that may have an impact on subject safety, wellbeing or the intent and conduction of the study;<br>4. History or presence of HIV, hepatitis B, hepatitis C. |

We plan to collect whole blood and plasma samples from two centers no less than 1100 patients (see Figure 1). The SNP array based whole-genome genotyping and total plasma protein N-glycosylation profiles will be measured for at least 384 participants. Expression profiles of approximately 40 disc specimens will be measured.

### MRI imaging, clinical and socio-demographic data

We require each patient from this study to have lumbar spine MRI images. MRI scanning will be performed on a 1.5 Tesla (or higher) tomograph and will include L1-S1 lumbar segments. T1 and T2 weighted images, acquired both in the axial and sagittal planes, will be obtained. Images will be used to assess the disc degeneration grade, presence and type of Modic changes and presence and severity of vertebral endplate defects. Disc degeneration grade will be estimated under the Pfirrmann classification from 1 to 5, with 1 corresponding to normal disc and 5 corresponding to the most severe degeneration [41]. Modic changes will be evaluated for each endplate (from the lower L1 vertebra to the upper S1 vertebra) on sagittal scans using T1WI and T2WI [42]. Vertebral endplate defects will be ranged from 1 to 6 according to the Rajasekaran classification [43]. Then, total endplate scores (TEPS) will be calculated as the sum of the endplate defect scores of both upper and lower endplates in each L1-S1 spinal segment. Additionally, height of the intervertebral discs and severity of osteophytes will be estimated based on Jarosz classification [44].

Clinical assessment will include demographic data (sex, age), self-reported ethnicity, height, weight, family status, physical activity, smoking, comorbidity, education level and job type.

All participants will also complete the following clinical questionnaires: visual analogue scale (VAS) [45] back and VAS leg estimating the intensity of back and leg pain correspondingly and the Oswestry disability index (ODI) [46,47] questionnaire.

### Sample collection, transporting and storing

Samples, collected from NRITO will be frozen at −40C (whole blood and plasma) or at −80C (disc samples) and then transferred to ICG for long-time storage at −80C. All biosamples from Priorov CITO will be frozen and stored on-site at −80C.

Blood and plasma sampling: from each patient 13 ml of peripheral venous blood will be collected into two BD Vacutainer 4 ml and 9 ml K2 EDTA tubes (or three 4 ml K2 EDTA tubes depending on availability). Tubes will be labeled with a unique patient’s code. During the first 30 min after blood collection, the 4 ml vacutainer with whole blood will be placed into the test tube rack and transported to the freezer to be stored at −40C. The other 9 ml blood tube will be used for plasma extraction according to the plasma extraction protocol (see Supplementary Materials 1). When the plasma will have been extracted, the tubes with plasma will also be labeled and placed into the freezer at −40C.

Eventually, all the tubes from NRITO will be transported to ICG in thermoboxes with cooling agents and then placed into freezers at −80C. Samples from Priorov CITO will be stored locally at −80C at once.

Plasma samples could be potentially used for cell-free DNA, glycomics, proteomics and metabolomics analyses.

Disc sampling: during the surgical procedure the resected fragments of intervertebral discs will be placed into sterile falcon 50 ml tubes that will be stored in the operating room till the end of intervention (no longer than 40 min). The falcons will be labeled with unique codes and will be transported to the laboratory in 5 min where the discs’ fragments will be put into liquid nitrogen.

Disc samples from NRITO will be transferred to ICG within thermoses with liquid nitrogen and then will be stored in freezers at −80C. Biospecimens obtained in Priorov CITO will be stored on the spot at −80C.

### Genotyping

DNA will be extracted according to the standard protocol with the help of Qiagen DNeasy Blood & Tissue Kit. Genotyping by at least 600,000 SNPs will be performed using whole-genome SNP-arrays of high coverage (tentatively, Illumina Infinium Global Screening Array will be used). Imputation procedure will be carried out using Haplotype Reference Consortium [48] or later reference panel.

### Total plasma proteins N-glycosylation profiling

The plasma glycans profiling will be performed using APTS glycan labelling kit (Genos) according to standard protocol [49].

### Total RNA profiling

The intraoperative material will be homogenized using TissueLyzer II homogenator (QUAGEN) and total RNA will be extracted and converted to cDNA using Kit for the isolation of total RNA and microRNA from cells and tissues (Biolabmix, Russia) and M-MuLV–RH First Strand cDNA Synthesis Kit (Biolabmix, Russia) respectively. The amount of extracted RNA and its quality will be estimated using Bioanalyzer 2100 (Agilent).

Total RNA sequencing will be performed using Illumina-HiSeq 4000 under PE-protocol. The read length up to 100 bp and sequencing coverage of 20M are expected. In total not less than 20 samples will be profiled.

### Duration of the project

Specimens collection and processing will be lasting for three years, unless the additional funding is obtained to expand the biobank. Samples will be stored for fifteen years after collection according to the storage protocol described above.

### Statistical data analysis

In this section we will describe designs of further studies that we plan to conduct based on the omics data generated from the samples of the biobank.

### Genome-wide association study of LDD and in-silico follow-up

We will perform genome-wide association study (GWAS) using quantitative scales of assessed disc degeneration status. The GWAS results will be used for replication of genome-wide significant findings from GWAS of LDD conducted in 2013 by Williams et al. [35]. Further, these two sets of GWAS will be meta-analysed to obtain the largest to date GWAS on LDD for European ancestry population. The results of meta-analysis will be used for in-silico follow-up functional annotation and prioritization of genes involved in LDD development.

### Functional analysis and data integration

GWAS meta-analysis results will be annotated in order to predict the probable effects of replicated SNPs on gene expression and disease development. Furthermore, we will highlight the molecular pathways, cell and tissue types most likely to be involved in DD pathogenesis. All this information will be used for gene prioritisation alongside with the results of analyses, based on colocalization analysis[50] and Mendelian randomization [51,52]. This methodology will be applied for causal inference between DD and gene expression profiles from different tissues, also including the transcriptomic data from intervertebral discs obtained in this study. A gene network of DDD regulation will be built and the key regulators will be revealed.

### Glycomics data analysis

The association of plasma N-glycans levels with disc degeneration status will be studied. Any obtained data may be helpful to use as dynamic glycan biomarkers for elucidating DD pathogenesis and may ultimately provide prognostic information.

### Transcriptomic data analysis

Patient group with available intraoperative disc material (see Figure 1) could be divided into “cases” or “degenerated disc” (grades 4-5 of disc degeneration according to Pfirrmann classification) and “controls” or “the healthy disc” (grades 1-3 of disc degeneration according to Pfirrmann classification) [43,53].

This division doesn’t influence the scheme of patient recruiting or sample collection but plays a role in the transcriptomic data analysis. The groups will include following ICD-10 codes: M51.1 “Thoracic, thoracolumbar and lumbosacral intervertebral disc disorders with radiculopathy” (cases); M51.3, M51.8 “Other thoracic, thoracolumbar and lumbosacral intervertebral disc degeneration” (controls).

Transcriptomic data obtained in this study will be used for detection of genes differentially expressed in discs between cases and controls. In short, the gene expression data will be mapped [54] on the reference genome and quality of the reads will be recalibrated (“Picard Toolkit.” 2019. Broad Institute, GitHub Repository. http://broadinstitute.github.io/picard/; Broad Institute). Subsequently, aiming to reveal the transcriptomic differences in intervertebral discs tissue between cases and controls, we will count the reads and identify differentially expressed genes (DEGs) [55]. At last, the revealed DEGs will be functionally annotated with Gene Ontology terms.

The data will also be used to identify expression quantitative trait loci (eQTLs). We will perform eQTL analysis of gene expression levels in intervertebral discs using methodology described in [56]. Resulting regional association summary statistics will be used for the gene prioritization.

### Data management and access

All data management and access procedures will be identical in both participating centers.

Each participant included in the study is assigned a unique code. Keys for these codes are saved in the locked storage of the internal hospital Electronic Data Capture (EDC) system, with access provided only for curators of the study. All clinical data, obtained from patients, are kept in the internal hospital EDC system, accessible only for authorized researchers filling data into it. Information on who entered the data into the clinical database is available for viewing. During the study, internal monitoring will be conducted to maintain the quality of the study in accordance with the GCP principles. Participant and specimen codes are transferred to ICG along with the specimens from both clinical centers in an anonymous way.

Information on the physical location of specimens is kept with limited access only for curators of the study. Genotypes, transcriptomic data and data produced during their processing will be stored on a local server in ICG.

To access the database, omcis data and other relevant information, projects should be submitted to the steering committee (contact the corresponding author) of the RuDDS.

## DISCUSSION

This study will be carried out in two different centers providing wider population coverage and more reliability to the sample storage, as data and specimens will be stored in two places. Participants will be recruited at unique medical centers (NRITO, Priorov CITO), that are the only centers in Russia specializing in studying DDD. The distinctive features of our clinical centers are: large constant flow of DDD patients, opportunity to conduct spine MRI and a highly-qualified team [57,58]. In the same way, our multidisciplinary research group, who will generate and analyze omics data, has a broad experience in studying back pain (one of the main manifestations of DDD) [59–61] and statistical and functional data analysis using integrative omics approaches [62,63].

Another notable advantage of the present project is availability of diverse and rare biological material that can be used for multiple profiling. Besides the whole blood samples, plasma and the intervertebral disc tissue obtained during the spine surgery will be collected. Presence of variable biological specimens for the same study participants will allow conducting integrative multi-omics analyses. Thus, not only genomes, but also transcriptomes, glycomes and other ‘-omes’ can be measured using these samples. To our knowledge it is the first study with multiple omics profiling of similar biological material. Current studies are limited by single profiling of one or two tissues [one - [32], [33]; two - [37], [31], [28], [29] or use omics datasets in open access, combining data from different cohorts [40]. The importance of the multi-omics approach is hard to overestimate, as it allows us to look at the disease from different points of view and level-up our understanding of the pathology.

One more substantial strength of our study is the assessment of disc degeneration grade by MRI scans because it is the most accurate and precise method of DD diagnostics among the others. In LDD studies based on plain radiography [64] or CT [65] phenotype definition is rather subjective as these approaches provide only indirect evidence of disc degeneration such as disc height loss and osteophytes. Similarly, the use of self-reported questionnaires for LDD identification [66] is not reliable enough. On the contrary, Pfirrmann grading system of disc degeneration used in the present study is based on MRI scans and estimates the main characteristics of DD by assessing the signal intensity and height of the intervertebral disc - the more dehydrated the disc is, the more severe degenerative changes are in it [41].

However, this study has some limitations. First, it should be noted that the expected sample size is modest in comparison with national country-level biobanks or large prospective cohorts. Sample size of the present study is four times less than one reported for meta-analysis of DDD studies reported by Williams et al. [35]. This limitation is explained by funding, establishing the recruitment period of three years. Nonetheless, the expected number of disc tissue samples (∼510) and transcriptome profiles of them (∼40) is comparable with other omics studies using disc specimens [proteome [33] - 7 cases / 7 controls; metabolome [39] - 60 cases / 21 controls; transcriptome [28] - 39 annulus fibrosus (AF) and 21 nucleus pulposus (NP) samples, [29] - 24 AF and 24 NP samples].

The next limitation of our study is imbalance between patients with “healthy disc” and “degenerated disc” in the group of patients subjected to surgery. Obviously, patients with “healthy discs” are subjected to spinal surgery under specific circumstances. Although, according to our protocol we still can expect some controls in this group (in total, the expected number of controls is 20). In groups not subjected to surgery we expect the ratio between cases and controls closer to the general population.

It should be noted that “healthy disc” is a conditional definition, not objective enough; however, the inter-observer agreement is quite high among different groups of researchers, therefore, Pfirrmann classification is considered to be highly reliable [67]. The main point here is that the existence of adult patients with perfectly healthy lumbar discs is questionable. To find such patients a huge large-scale exploratory study is needed. Nevertheless, according to a population-based study there are no adults with lumbar discs containing no degenerative changes [13].

## ETHICS AND DISSEMINATION

The study will be performed according to the Helsinki Declaration; the study protocol was approved by the Local Ethical Committee of NRITO (№034/20 dated 02 Oct 2020) and by the Local Ethical Committee of Priorov CITO (№1/21 dated 25 Feb 2021). Preliminary and final results will be presented in peer-reviewed journals, at national and international congresses.

## AUTHORS’ CONTRIBUTIONS

ONL, EEE, AVK, YSA and YAT contributed to the study concept and design of the study protocol. ONL, EEE, TSG, YAT contributed to the development of specimen collection, storage and profiling protocols. ONL, AVP and AVK contributed to the setting of the logistics of working with hospitals and ethical committee approval. ONL, EEE and YAT drafted the first version of the manuscript. All authors critically reviewed and approved the final manuscript.

## COMPETING INTERESTS STATEMENT

YSA is a co-owner of Maatschap PolyOmica and PolyKnomics BV, private organizations, providing services, research and development in the field of quantitative and computational genomics.

### Data statement section

Prepublication history for this paper is available online. To view these files, please visit the journal online.

