## Supplementary Materials 1 for "Protocol for disease-oriented Russian disc degeneration study (RuDDS) biobank facilitating functional omics studies of lumbar disc degeneration"

### STANDARD OPERATING PROCEDURE FOR PLASMA COLLECTION

**General conditions**

1. Safety and personal protective equipment

The following personal protective equipment (PPE) should be worn: laboratory coat, laboratory gloves, safety mask.

2. Environmental conditions: biological sterility

Plasma isolation is conducted in the laminar flow cabinet (class II microbiological cabinet - biohazard according to EN12469 standard). The work surface and equipment are cleaned with water and then with 70% ethanol at the end of the procedure.

3. Temperature range: all procedures are performed at room temperature (23 ± 3˚C)

4. Laboratory equipment

- Laminar flow cabinet Mars Safety Class 2
- Freezer (-40°C or -80°C) SANYO Biomedical freezer MDF-U33V
- Eppendorf Centrifuge 5810 R
- Rainin 100-1200 µL Multi Channel pipette
- Pipette tips Vertex 1250 µL Filtered (4347NSF 18262)
- Improvacuter K2 EDTA 4ml, 9 ml
- Vacuette tubes 3ml no additive
- TR Safe Tube 1.5ml

**Plasma collection procedure**

Steps 2, 4 and 6 are performed in the laminar flow cabinet

**Step 1: Collect blood samples from participants**

From each participant vacuum EDTA blood collection tubes (two tubes 4 ml each or one tube 9 ml) are required. All the tubes must be labeled with a unique patient ID.

**Step 2: Incubation of blood samples at room temperature** (performed in the laminar flow cabinet)

Leave the tubes resting in upright position at room temperature for an hour.

**Step 3: First centrifugation of the blood samples**

• Without disturbing blood fractions, gently place the vacuum tubes into the centrifuge, balance the samples.

• Centrifuge the tubes at 1620 g for 10 minutes. Note: Do NOT use brake to stop the centrifuge!

**Step 4: Transfer Plasma to Clean Vacuum Tubes** (performed in the laminar flow cabinet)

• Label clean vacuum tubes with the unique patient ID.

• Without breaking the plasma/formed elements interface, immediately transfer the plasma to a clean vacuum tube using filter tips. At least 4 ml of plasma is taken from tubes prepared at Step 1. IMPORTANT: change tips after each sample!

**Step 5: Second centrifugation of the blood (plasma) samples**

• Place the tubes containing plasma in centrifuge, balance the samples.

• Centrifuge the tubes at 2700 g for 10 minutes. Note: Do NOT use brake to stop the centrifuge!

**Step 6: Filling the plasma into cryotubes** (performed in the laminar flow cabinet)

• Label cryotubes with a unique patient ID.

• Label three cryotubes containing plasma from the same patient with 1, 2 and 3.

• Transfer the plasma to 1 ml cryotubes. Thus, 3 aliquots of 1 ml plasma will be obtained from each patient. IMPORTANT: change tips after each sample!

**Step 7: Plasma storage**

Transfer the cryotubes containing processed plasma to a rack and store in a freezer at - 80°C (or at - 40°C).

**Step 8: Clean the work surface**

At the end of the work, wipe the laminar work surface and all used automatic dispensers first with water and then with 70% ethanol.
